# Laboratory validation of a simplified DNA extraction protocol followed by a portable qPCR detection of *M. tuberculosis* DNA suitable for point of care settings

**DOI:** 10.1101/2024.04.05.24305384

**Authors:** Tainá dos Santos Soares, Graziele Lima Bello, Ianca Moraes dos Santos Petry, Maria Rita Castilho Nicola, Larissa Vitoria da Silva, Regina Bones Barcellos, Joana Morez Silvestri, Maria Lucia Rossetti, Alexandre Dias Tavares Costa

## Abstract

Tuberculosis, caused by *Mycobacterium tuberculosis*, is a treatable and curable disease, and yet remains one of the leading causes of death worldwide. Diagnosis is essential to reducing the number of cases and starting treatment, but costly tests and equipments that require complex infrastructure hamper their widespread use as a tool to contain the disease in vulnerable populations as well countries lacking resources. Therefore, it becomes necessary to develop new technological approaches to molecular methods as well as screening tests that can be rapidly conducted among people presenting to a health facility to differentiate those who should have further diagnostic evaluation for TB from those who should undergo further investigation for non-TB diagnoses. The present study aimed to evaluate two experimental DNA extraction methods from clinical samples (FTA card versus sonication) followed by analysis in a portable qPCR instrument (the Q3-plus). The FTA card-based protocol showed 100% sensitivity and specificity, while the sonication protocol showed 80% sensitivity and 89% specificity when compared to the traditional gold standard culture. The portable protocol, comprised by the FTA card method and the portable instrument Q3-Plus, showed sensitivity and specificity of 92% and 61%, respectively, when compared to culture, and 75% and 81%, respectively, when compared to the standard TB case classification. The ROC curve showed an AUC of 0.78 (p<0.001) for the portable protocol and 0.93 (p<0.001) for the GeneXpert MTB/RIF. The limit of detection (LOD) for *Mycobacterium tuberculosis* (H37Rv strain) detection in spiked samples obtained using the portable protocol (FTA card and Q3-Plus) was 19.3 CFU/mL. As an added benefit, using the FTA card facilitates sample handling, transport, and storage. It is concluded that the use of the FTA card protocol and the Q3-Plus yields similar sensitivity and specificity as the gold standard diagnostic tests and case classification. We suggest that the platform is suitable to use as a point of care tool, assisting in the screening of tuberculosis in hard-to-reach or resource-limited areas.

## Introduction

Tuberculosis (TB) is one of the deadliest diseases in the world. In 2021, 10.6 million people developed active TB and 1.6 million people died [1]. In Brazil, in 2021, 5.072 people died and 78.057 were diagnosed positive in 2022 [2]. Traditional TB diagnosis occurs through sputum smear microscopy, an inexpensive and practical method, but with limited specificity and sensitivity. Mycobacterial growth culture is considered the gold standard method and, although it takes too long to produce results (usually around 3 to 4 weeks), it has the advantage of achieving high sensitivity [3,4].

Molecular tests emergence overcame some of the limitations presented by traditional tests, providing highly specific and sensitive diagnosis in just a few hours. Real-time PCR (qPCR) allows the amplification and quantification of nucleic acids and is currently the most used molecular technique for diseases diagnosis [5,6]. In the Brazilian Unified Health System (SUS), the GeneXpert MTB/RIF^®^ assay (Cepheid, USA) is the test of choice, providing results in up to 2 hours, and a bonus of being recommended by WHO since 2010. This test amplifies specific targets of *Mycobacterium tuberculosis (Mtb)* genome and the rifampin resistance determining region (RRDR) in *rpo*B gene [3,5,6]. However, GeneXpert MTB/RIF assay also has limitations, such as false-negative results, which were partially solved in the tests’ newer versions, such as GeneXpert MTB Ultra^®^ [6,7,8]. The implementation of the GeneXpert MTB/RIF assay increased the number of positive diagnoses, but did not improve global case detection rates, as the equipment has high infrastructure requirements and costs, making it not viable for resource-limited areas where the TB burden is higher [3]. An alternative to increase the number of diagnoses is active search, using effective and accessible diagnostic tools with a rapid response time that can perform in resource-limited settings [4].

The implementation of tests where patients are treated (“point of care”, or POC) allows for the referral of only confirmed cases to health centers [5, 6]. However, alternative screening methods are still needed to facilitate patients access to these technologies, reducing the initial barrier to obtain some medical evaluation and increasing the motivation of individuals with a higher probability of developing TB [7].

One of the challenges for molecular and portable screening tests is defining a process to obtain DNA that is simple and fast and does not require dangerous chemicals, complex biosafety infrastructure, manipulation, or storage [5,7]. The method of choice must be fast, simple, low cost, and still provide a good quantity and quality (i.e. purity) of material. The extraction technique to be used must vary according to the sample type and the downstream applications, with varying steps of cell disruption, removal of lipids, proteins, and other nucleic acids, as well as purification and concentration of the targeted nucleic acids. For example, rupture of the cell wall can be performed mechanically, enzymatically, chemically, or by combinations of these techniques [9,10,11]. There are many methods of DNA extraction available as commercial kits for application in molecular analysis. Some laboratories, on the other hand, prefer to use in-house methods to reduce costs [12,13]. Among the in-house methods already standardized, sonication deserves special consideration. This method is considered simple and very effective for breaking cell walls, without the need of chemical reagents, enzymes, proteins, or substances that can compromise the integrity and detection of the sample’s DNA [9,10]. Alternatively, simple protocols using the FTA Elute Micro card (Whatman, USA) have been published [14,15,16]. Detergents are embedded in these FTA cards, making them multifunctional: in addition to being a transport and storage medium, detergents also help to solubilize cell membranes, thus releasing DNA in the extraction processes, which then binds to the cellulose matrix and is easily eluted with aqueous solutions whenever necessary. The sample attached to this card is less susceptible to contamination, keeping the material intact for years without the need for costly conservation strategies [9,11,14].

WHO recognizes the need for more sensitive and specific tests to improve the early diagnosis of TB, such as the development of portable POC devices, which stand out for their simplicity, accessibility, and portability [1,5,7]. Therefore, our study aimed to evaluate two DNA extraction protocols for the detection of *Mtb* from sputum, one sonication-based and the second FTA card-based [15]. The chosen protocol was further validated using a portable qPCR system, the Q3-Plus [17], in parallel to the routine diagnostic testing using the GeneXpert MTB/RIF assay and culture.

## Methods

### Samples origin

We used 127 sputum samples from patients over 18 years old, seeking treatment at the Department of Tisiology and Leprosy of the Health Department at the University Hospital of the Brazilian Lutheran Universtiy at Canoas (ULBRA), in the municipality of Canoas (south of Brazil), from October 1^st^ 2018 to September 30^th^ 2022. Written informed consent was obtained from all participants, and no minors were included. We used 29 samples, divided into two aliquots, totaling 58 samples to test protocol 1 and protocol 2 and 98 samples were used for validation portable instrument Q3-Plus. Samples with a minimum volume of 500 μL were characterized for presence or absence of *Mtb* by GeneXpert MTB/RIF and culture (Bactec MGIT). After routine characterization, samples were sent to the Molecular Biology Laboratory at the Lutheran University of Brazil (ULBRA), where they were processed and submitted to molecular tests objective of the present study.

### Standard clinical classification of tuberculosis cases (“case TB”)

WHO defines a tuberculosis positive case as definitive when *Mtb* is identified in a patient’s sample by either culture or a molecular assay [18]. In countries that do not have the laboratory capacity to routinely identify *Mtb*, a pulmonary case with one or more initial sputum smear testing positive for acid-fast bacilli (AFB) is also considered a “definite” case, if there is a functional external quality assurance (EQA) system with blind rechecking. In this study, we classified all samples with positive culture or positive GeneXpert assay as positive TB cases according to SINAN (Brazilian Notifiable Diseases Information System) and the TB Site (Tuberculosis Special Treatment Information System) [18]. Therefore, the diagnostic technique for these samples will be referred as “clinical case classification (or simply “case TB”)”.

### DNA extraction protocols

#### Sonication-based

Sputum samples were decontaminated with 4% sodium hydroxide (NaOH) in PBS (phosphate-buffered saline). A total of 500 μL of sputum was transferred to a 1.5 mL microtube and 4% NaOH was added, followed by vortex homogenization, and incubation at 37 °C for 15 min. Subsequently, it was centrifuged 3000 g for 15 min and the supernatant was discarded. The pellet was resuspended in 500 µL of PBS and homogenized on a vortex. Next, the tube was incubated at 95 °C for 20 min. After this step, it was placed in the sonication bath for 15 min, and then centrifuged for 5 min at 3000 g. Finally, 100 μL of the supernatant (containing the DNA) was transferred to a new microtube and frozen at −20 °C until use [19,20].

#### FTA card-based

DNA extraction with FTA^®^ Elute Micro card (Whatman, USA) was performed as described by Ali *et al* 2020 [15], with minor modifications. Briefly, an aliquot of the sample was mixed with a solution containing 6 M guanidine isothiocyanate and 0.5 M EDTA (ratio of 400 μL denaturing solution per 1000 μL sample) and 20 μl of Proteinase K (25 mg/ml, Roche Diagnostics, Germany). Next, the sample was vigorously shaken in vortex for 20-30 seconds. The mixture was evenly applied to a FTA^®^ Elute Micro card using a plastic Pasteur pipette, and airdried for at least 1 hour at room temperature. From this point on, the card can be used directly for further processing or for sample storage. For DNA extraction, a 6 mm diameter disc was punched out and placed in a 1.5 mL microtube. TE buffer pH 8.0 (500 μL) was added to the disc-containing tube which was then vortexed and incubated at 95°C for 5 min. The tube was then centrifuged for 1 minute at maximum speed, and 50-100 μL of the supernatant was transferred to a new tube and stored at -20°C until qPCR amplification [15].

### Real-time PCR

DNA amplification and *M. tuberculosis* detection was performed by real-time PCR (qPCR), in a Step One Real Time PCR System (AB Applied Biosystems, USA). Primers for detecting the *IS6110* genomic marker (Forward 5’-CAGGACCACGATCGCTGAT-3’ and reverse 5’-TGCTGGTGGTCCGAAGC-3’) and the probe (5’-FAM-TCCCGCCGATCTCG-HQI-3’) were used. Each reaction had a final volume of 10 μL, containing 0.5 μL oligomix (20X primer and probe solution), 3.5 μL PCR mix (Kapa Probe Fast qPCR Mastermix), 3.5 μL ultrapure water and 2.5 μL extracted DNA. The reference strain H37RV (10 ng/μL) was used as a positive control (PC) and ultrapure water was used as a negative control (NC). Amplification conditions were as follows: 50°C for 2 minutes, followed by 10 minutes at 95°C, and 95°C for 15 and annealing and amplification at 60°C for 1-minute seconds for a total of 40 cycles. [21]

DNA amplification and detection of the *IS6110 M. tuberculosis* genomic marker using the portable Q3-Plus instrument (ST Microelectronics, Italy) was performed using the same reagents described above, but for a final reaction volume of 5 μL. The reaction contained 0.25 μl of *IS6110* oligomix, 2.5 μl of qPCR master mix, 0.75 μl of ultrapure water, and 1 μl of extracted DNA. The amplification conditions were as follows: 95°C for 10 minutes, followed by 45 cycles of 95°C for 15 seconds and 60°C for 60 seconds. The optical parameters for the FAM channel in the Q3-Plus system were exposure time of 1 second, LED power of 3, and analog gain of 15, while for the HEX channel the optical parameters were exposure time of 2 seconds, LED power of 10, and analog gain of 15. The reaction was supplemented with 0.5 μl oligonucleotides for detection of the human 18S rRNA gene (Forward 5’ TGCGAATGGCTCATTAAATC 3’, Reverse 5’ CGTCGGCATGTATTAGCTCT, and HEX-probe TGGTTCCTTTGGTCGCTCGCT-BHQ1), which was used as an internal control [15]. In both instruments, baseline and threshold were set to automatic.

### Limit of Detection (LOD_95%_): Colony forming units in parallel to qPCR

Reference strain *Mtb* H37Rv colonies cultivated in the Ogawa-Kudoh medium were collected and homogenized with glass beads. The turbidity of the bacterial solution was compared to the turbidity of the McFarland number 1 standard. Subsequently, ten-fold serial dilutions were performed from the concentrated cell suspension, and tubes containing 500 μL of mucin suspensions (20% v/w) were spiked with 30 μL of each dilution. The whole volume was equally distributed onto individual FTA Elute Micro cards and the simplified DNA extraction protocol was performed (“FTA card-based protocol” above). This procedure was performed in duplicate for each cell suspension concentration. The same procedure was performed in parallel and spiked samples were plated in petri dishes containing 7H10/OADC medium, in duplicates. Colonies were counted using a semi-quantitative scale, according to the current Brazilian standards [19].

### Statistical Analysis

Reactions on the Step One instrument was performed in triplicates, and reactions on the Q3-Plus instrument were performed in duplicates (optimization protocols) or quadruplicates (patient samples). The Cohen’s kappa coefficient was calculated between the Q3-Plus or Step One results obtained with the FTA card-based protocol and the results obtained with GeneXpert or culture, using these latter methods as the gold standard with a 95% CI (confidence interval). The Kappa (K) agreement force was interpreted as follows: Strength of Agreement < 0.00 (Poor), 0.00-0.20 (Slight), 0.21-0.40 (Fair), 0.41-0.60 (Moderate), 0.61-0.80 (Substantial) and 0.81-1.00 Almost Perfect) [12]. Sensitivity and specificity were evaluated about the tuberculosis case definition by the attending physician. Student t-test with a significance level of 0.05 was used to evaluate the difference between results obtained with the Step One and the Q3-Plus system. The LOD_95%_ for the DNA extraction protocols was estimated by qPCR in parallel to colony growth using a Probit regression.

All data were analyzed using Statistical Package for the Social Sciences (SPSS), version 21.0.

All relevant data are within the manuscript and its Supporting Information files.

## Results

### Sample analysis

Sputum samples from 29 patients were characterized by culture and GeneXpert MTB/RIF test before analysis by the two experimental DNA extraction protocols. Data from GeneXpert MTB/RIF test revealed that 14/29 (48.2%) were positive and 15/29 (51.7%) were negative samples, while culture analysis showed that 11/29 (38%) were positive and 18/29 (62%) were negative samples.

Samples were also aliquoted for analysis by both experimental extraction methods (Sonication- and FTA card-based). The presence of *Mtb* was confirmed in the final eluate of each protocol by qPCR (Figure 1). The sonication protocol showed that 10/29 (34,4%) were positive and 19/29 (65,5%) were negative samples, while the FTA card protocol revealed that 11/29 (38%) were positive and 18/29 (62%) were negative samples. Overall, 24/29 samples agreed as positive or negative between all four techniques. These results are also shown as a direct comparison of the results obtained by each experimental protocol per sample (Figure 2, Table 1). The data show that Cts obtained by the sonication protocol are lower than those obtained by the FTA card protocol by an average of – 2.5 Cts, but with higher variation (26.7 ± 7.50 versus 29.2 ± 4.52). However, the FTA card protocol yielded more positive samples than sonication.

**Table 1.** Cts corresponding to the detection of *IS6110* in DNA extracted by each protocol (sonication or FTA card) from TB-confirmed clinical samples.

| Sample | Sonication<br>(Ct) | FTA card<br>(Ct) | GeneXpert<br>MTB/RIF | Culture |
| --- | --- | --- | --- | --- |
| 1 | - | 35.83 | - | - |
| 2 | - | - | - | - |
| 3 | 37.70 | - | - | - |
| 4 | 21.45 | 24.90 | + | + |
| 5 | - | - | - | - |
| 6 | - | - | - | - |
| 7 | - | - | - | - |
| 8 | - | 35.73 | - | - |
| 9 | - | - | - | - |
| 10 | - | 36.95 | - | - |
| 11 | 36.75 | - | - | - |
| 12 | - | - | - | - |
| 13 | - | - | - | - |
| 14 | - | - | - | - |
| 15 | 31.66 | - | - | - |
| 16 | 38.42 | 35.16 | + | - |
| 17 | 19.24 | 25.98 | + | + |
| 18 | 18.57 | 26.32 | + | + |
| 19 | 25.21 | 28.84 | + | + |
| 20 | - | 30.80 | + | + |
| 21 | 33.97 | - | + | - |
| 22 | 30.54 | - | + | - |
| 23 | 20.19 | 27.37 | + | + |
| 24 | - | - | - | - |
| 25 | 18.34 | 22.85 | + | + |
| 26 | 24.28 | 26.14 | + | + |
| 27 | 17.34 | 30.26 | + | + |
| 28 | - | 24.45 | + | + |
| 29 | 20.39 | 26.40 | + | + |

When the results obtained by both experimental protocols were compared with the GeneXpert MTB/RIF results, the FTA card protocol detected 11 *Mtb* samples with GeneXpert assay detecting an additional 3 samples (#16, #21, and #22). The sonication protocol detected 9 samples with the GeneXpert assay detecting 4 more samples (#15, #20, #21, and #28). Compared to the culture, two samples (#20 and #28) presented a false negative result and one (#22) presented a false positive result by the sonication protocol, while the FTA card protocol showed 100% agreement with the culture (Table 2).

Coheńs agreement between the results of the experimental protocols (sonication versus FTA card) showed a kappa index of 0.7 (95% CI: 0.69 -0.79; “substantial”). The same level of agreement was found when comparing the sonication protocol versus culture (kappa = 0.7; 95% CI: 0.69 - 0.79). Interestingly, when the FTA card protocol was compared to culture, it showed a kappa index of 1.0 (95% CI: 0.80 – 0.99), higher than GeneXpert’s index of 0.7, thus suggesting a better performance (Table 2). The relationship between the results obtained by each protocol is shown in the Venn diagram in Figure 3. One sample was detected solely by sonication extraction and GeneXpert MTB/RIF, two samples were detected only by GeneXpert MTB/RIF, and two samples were detected simultaneously by GeneXpert MTB/RIF, culture, and FTA card extraction protocol, while nine samples were detected by all protocols. Sensitivity (SE) and specificity (SP) of the methods were determined in comparison with culture (Table 3). The protocol using extraction by sonication presented a SE of 80% (95% CI: 63 - 96) and SP of 89% (95% CI: 76 - 102), while the protocol using FTA card presented 100% SE and SP (95% CI: 98 - 101). The gold standard molecular test GeneXpert MTB/RIF displayed an SE of 100% (95% CI: 98 - 101) and SP of 79% (95% CI: 60 - 95).

**Figure 1.**
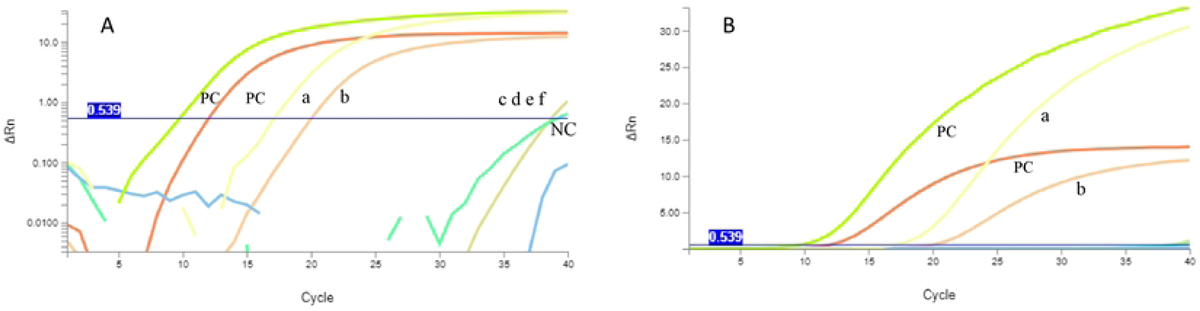
Representative curves of qPCR detection of the *Mtb* by the Step One (panel A) or the Q3-Plus (panel B) instruments. Representative curves for qPCR detection of the *Mtb* genomic marker *IS*6110 in samples processed by the sonication or the FTA card experimental protocols. “PC” and “NC” represent the positive control (25 ng/μL of DNA extracted from H37Rv MTB cells) and the negative control (TE pH 8.0). Traces “a” and “b” were obtained with sonication and FTA card protocols using sample #4, traces “c” and “d” were obtained with sonication and FTA card protocols using sample #13, and traces “e” and “f” obtained with sonication and FTA card protocols using sample #14, respectively. Traces are representative of at least three independent experiments for each protocol.

**Table 2.** Sensitivity, specificity, and kappa index of results from both experimental extraction protocols compared to GeneXpert MTB/RIF and culture.

|  |  | GeneXpert<br>MTB/RIF |  | Culture |  | Comparison to culture |  |  |
| --- | --- | --- | --- | --- | --- | --- | --- | --- |
|  |  | Positive | Negative | Positive | Negative | Sensitivity | Specificity | Kappa |
| Sonication | Positive | 10 | 0 | 9 | 1 | 80% | 89% | 0,7 |
|  | Negative | 4 | 15 | 2 | 17 | (95% CI:<br>63-96) | (95% CI:<br>76-102) | (95% CI:<br>0,6-0,79) |
| FTA card | Positive | 11 | 0 | 11 | 0 | 100% | 100% | 1,0 |
|  | Negative | 3 | 15 | 0 | 18 | (95% CI:<br>98-101) | (95% IC:<br>98-101) | (95% CI:<br>0,98-1,01) |
| GeneXpert<br>MTB/RIF | Positive |  |  | 11 | 3 | 100% | 79% | 0,7 |
|  | Negative |  |  | 0 | 15 | (95% CI:<br>98-101) | (95% CI:<br>60-95) | (95% CI:<br>0,6-0,79) |

**Figure 2.**
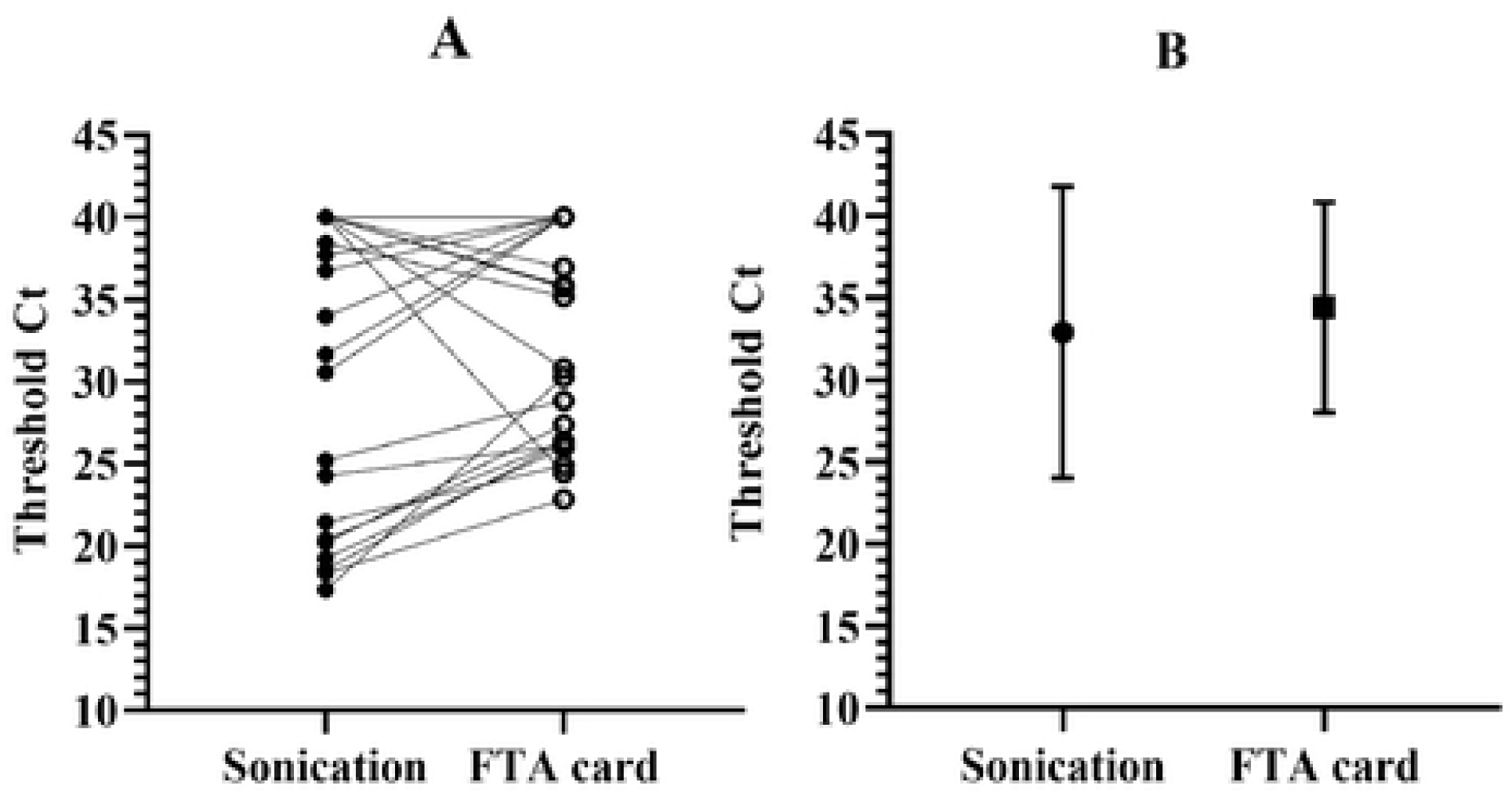
Comparison of threshold cycles between the sonication and card FTA qPCR for detection of *IS6110*. Panel A shows the same samples processed by each experimental protocol. qPCR was performed in the standard StepOne instrument. Data is shown as the average threshold Ct for each sample, which was tested in duplicate. Panel B shows mean ± SD for each experimental protocol, averaging all samples.

**Figure 3.**
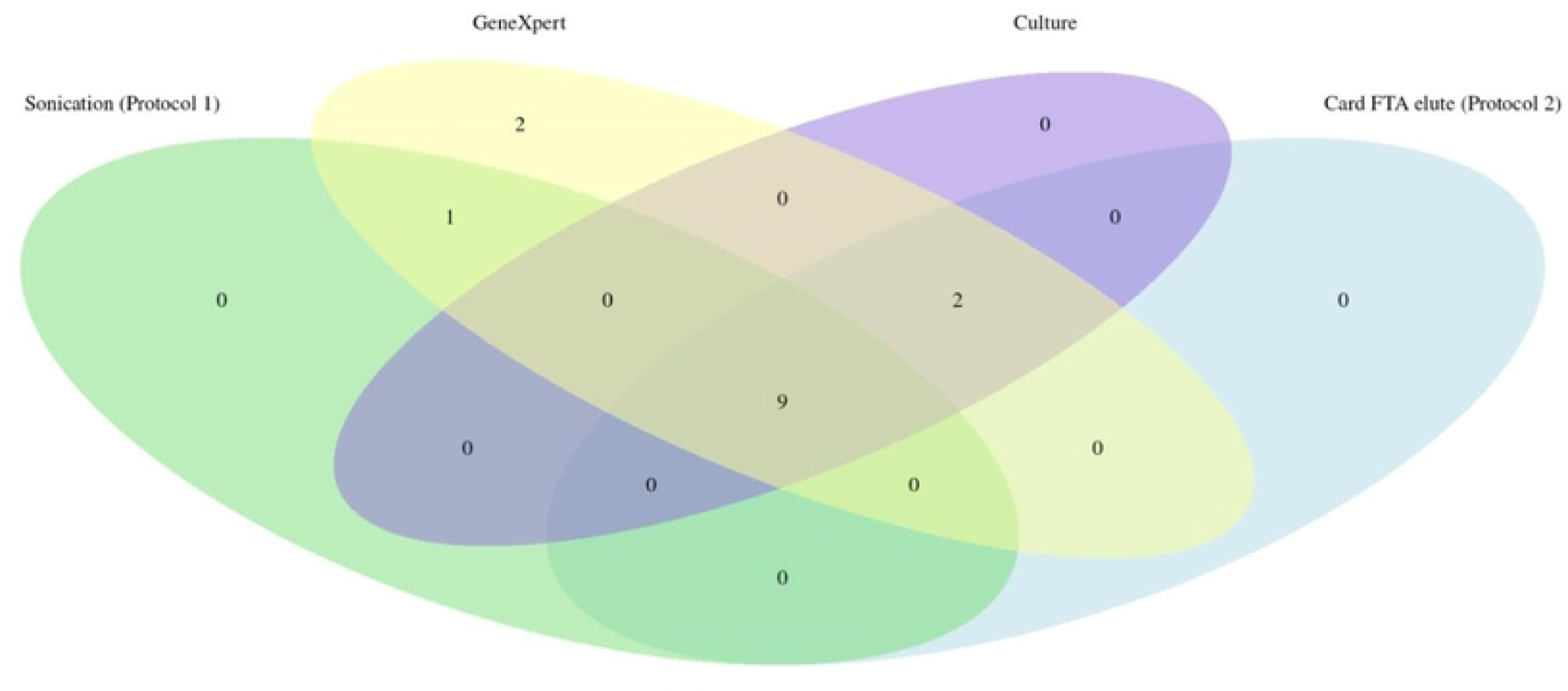
Venn diagram showing the number of samples detected by each technique. Numbers in overlapping areas indicate that the techniques agreed with the classification, whether the sample was negative or positive. All four techniques equally detected the presence of MTB DNA in 9 samples, while 2 were detected by FTA card protocol, culture, and GeneXpert assay. One sample was detected only by the GeneXpert assay and sonication, and 2 were solely detected by the GeneXpert assay.

Since the FTA card protocol detected more positive samples than the sonication protocol and yielded better sensitivity, specificity, and kappa index, it was chosen for the remainder of the present study on a simplified and portable DNA extraction method.

### Evaluation of qPCR with intact *Mtb* cells and colony-forming unit counting

Starting from a *Mtb* suspension corresponding to turbidity 1 on the McFarland scale, colony growth could be observed up to the fifth 1:10 dilution. Counting could be performed up to the fourth and fifth dilutions, yielding an average of 9.5 ± 6.3 and 0.5 ± 0.7 CFU/ml, respectively (Table 3). DNA extracted from each dilution by the simplified FTA card-based protocol was used for qPCR detection of *M. tuberculosis* DNA in the portable Q3-Plus system (Figure 4, panel A). The qPCR efficiency was determined to be 118% (slope -2.87 and R2 = 0.95) for Step One, and 216% (slope -1.96 and R^2^ = 0.99) for the Q3-Plus (Table 3, Supplemental Table S1). The FTA card protocol presented an apparent LOD_95%_ of 19.3 CFU/mL of MTB when the portable Q3-Plus instrument was used (Figure 4, panel B). We were not able to calculate the LOD_95%_ when the StepOne instrument was used because all replicates were positive.

**Table 3.** Colony forming unit and IS6110 detection by two qPCR instruments using 1:10 dilutions of a *M. tuberculosis* cell suspension.

| Dilution | Average (CFU/mL) | Step One (± SD) | Q3 Plus (± SD) |
| --- | --- | --- | --- |
| 1:10 | countless | $23.7 \pm 0.9$ | $25.7 \pm 2.1$ |
| 1:100 | $175 \pm 35.3$ | $29.4 \pm 0.7$ | $27.9 \pm 0.4$ |
| 1:1.000 | $35.5 \pm 6.3$ | $31.5 \pm 0.1$ | $29.2 \pm 0.8$ |
| 1:10.000 | $9.5 \pm 6.3$ | $33.3 \pm 1.1$ | $32.1 \pm 0.7$ |
| 1:100.000 | $0.5 \pm 0.7$ | $36.1 \pm 1.1$ | $36.5 \pm 1.4$ |

**Figure 4.**
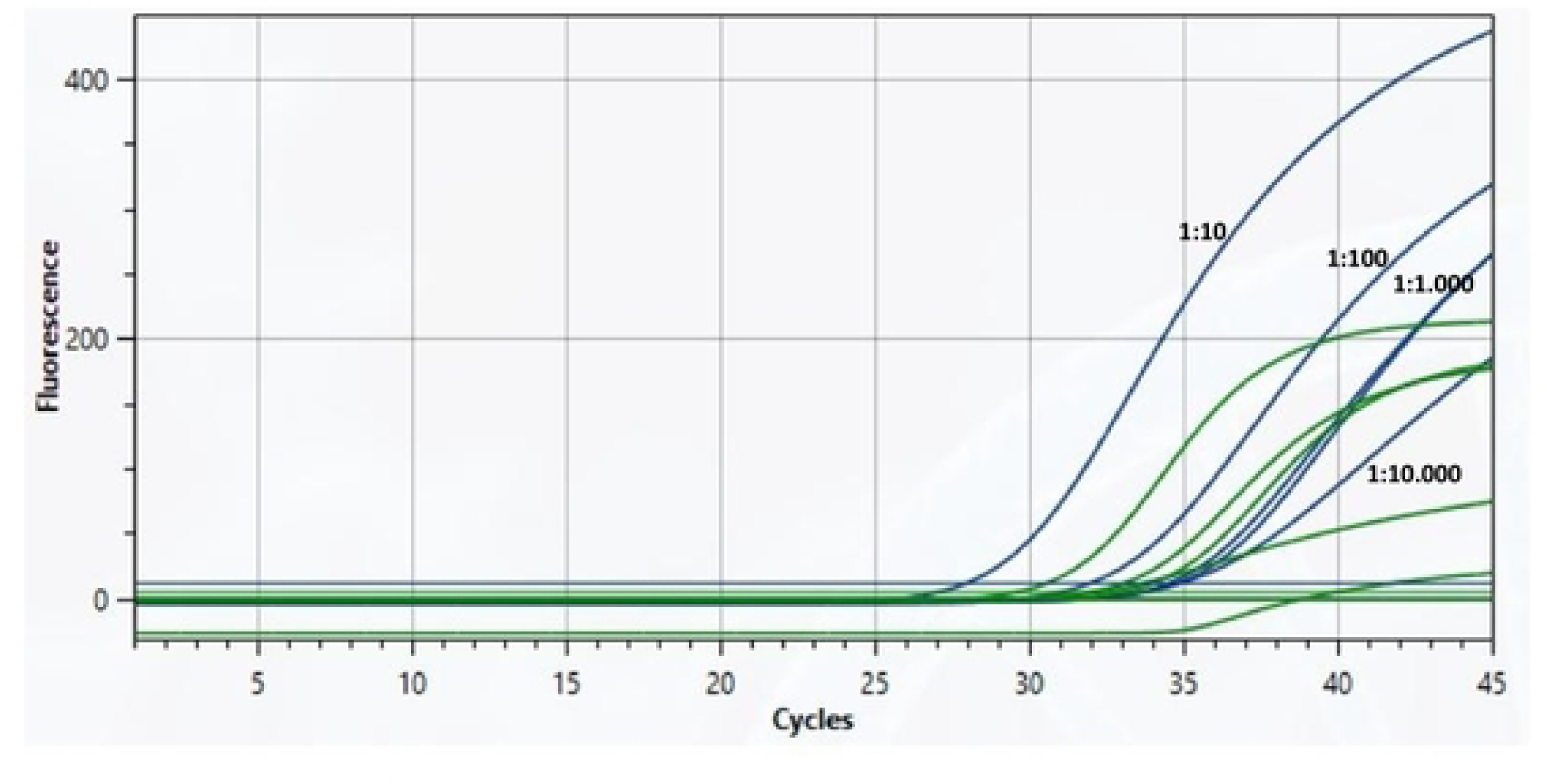
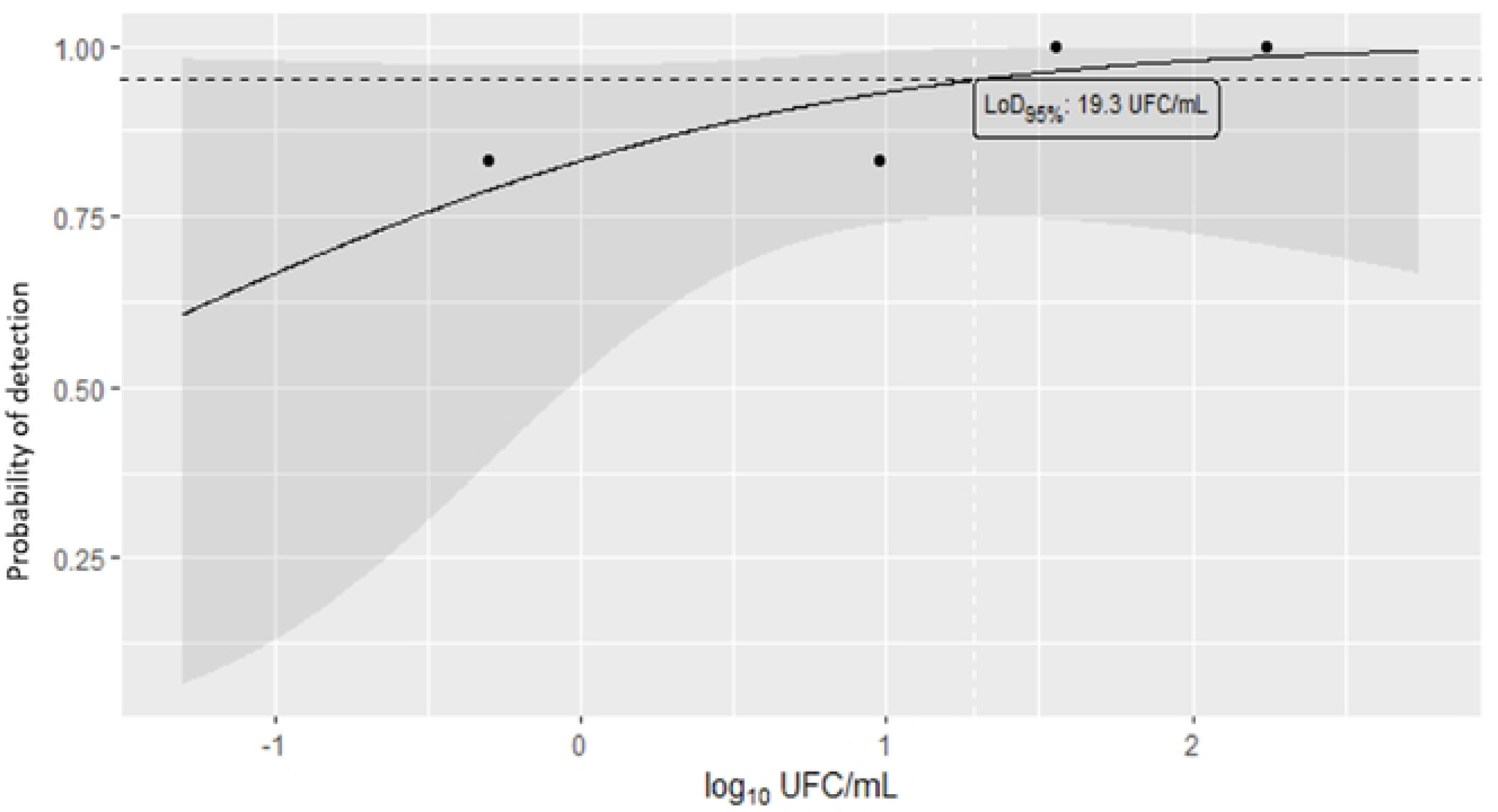
Representative traces for the detection of *Mtb DNA* by the portable qPCR instrument Q3-Plus and its calculated limit of detection probability as measured by colony forming units (CFU). Panel A. Shown are traces for the detection of the *Mtb* IS6110 genomic marker in the Q3-Plus qPCR system, using DNA extracted by the FTA card experimental protocol from dilutions 1:10, 1:100, 1:1.000, and 1:10.000 dilutions. Such traces were used to calculate the data shown in Table 3. Each sample was run in duplicate—panel B. A Probit regression analysis was performed on the probability of detection of each *M. tuberculosis* concentration by the Q3-Plus system, as shown in Table 3, yielding a LOD_95%_ of 19.5 CFU/mL.

Next, we validated the completed portable solution (DNA extraction + qPCR) using 98 samples in parallel to the routine diagnostic service. Sputum samples were processed by the FTA card protocol and evaluated for the presence of *Mtb* DNA in the portable and in benchtop instruments. Figure 5 shows representative curves of IS6110 and 18S rRNA detection as detected by Q3-Plus using DNA extracted from sample #34. Of the 98 samples, 34 (34,6%) were positive for *Mtb* DNA in the Q3-Plus and 45 (45,9%) were positive on GeneXpert MTB/RIF, with 27 (27,5%) samples being detected by both instruments.

**Figure 5.**
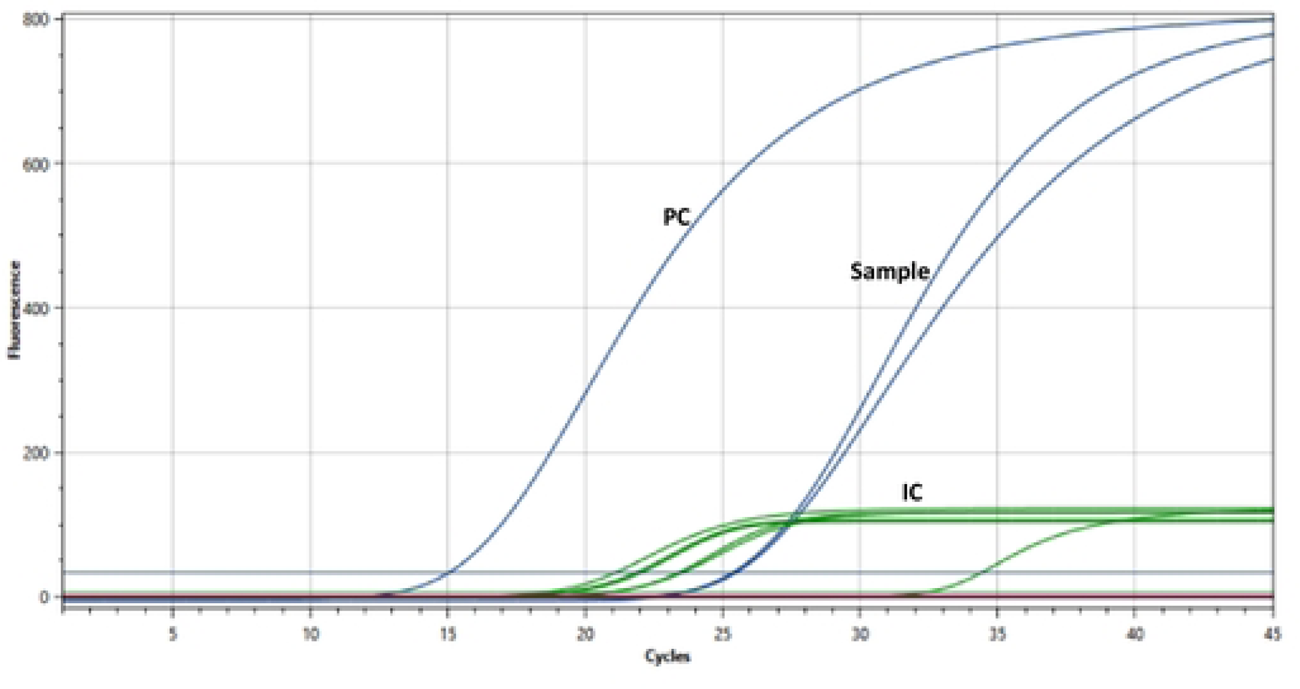
Representative traces of qPCR detection of *M. tuberculosis* IS6110 and human 18S rRNA obtained from a human sample. Representative traces of qPCR detection of the *Mtb* genomic marker *IS6110* (blue lines) and the human 18S rRNA (green lines, “IC”) for sample #34 (“sample”) as well as the positive control H37Rv DNA (10 ng/μL, “PC”). Each sample was tested in duplicate.

We then produced a ROC curve comparing the results obtained by the portable platform (FTA card protocol followed by qPCR in the Q3-Plus) and the GeneXpert MTB/RIF system against the classification of samples by TB case, the WHO reference standard method. The plots yielded an area under the curve (AUC) of 0.78 for the portable testing solution and 0.93 for the GeneXpert MTB/RIF assay. As p<0.001, it implies that these results are statistically significant.

**Figure 6.**
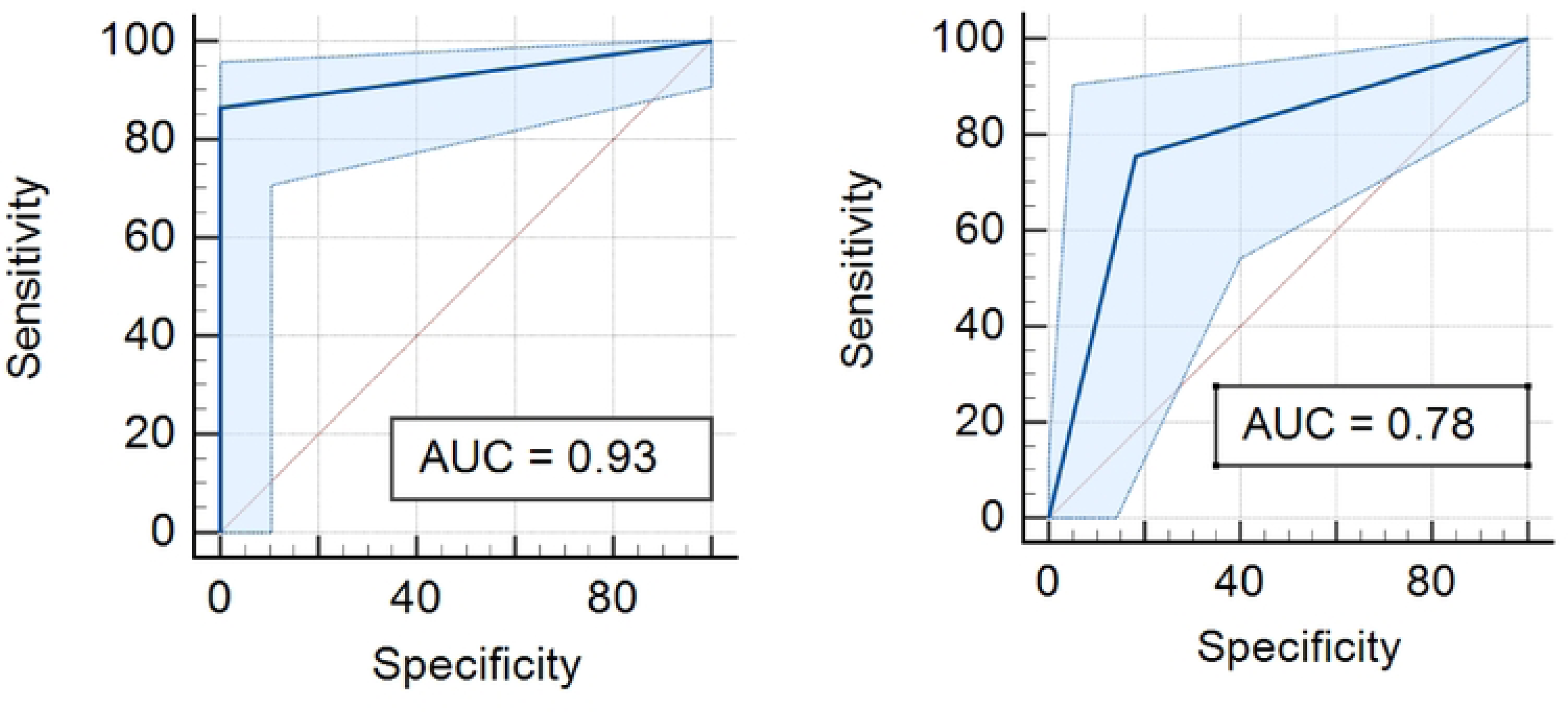
ROC curve for the detection of *IS6110 Mtb* marker using the Q3-Plus and FTA card protocol or the GeneXpert MTB/RIF system versus culture. Results obtained after the analyses of all samples by both molecular methods are plotted against the sample classification as shown by culture, the gold standard method. Diagonal segments were produced by bonds.

The Q3 Plus provides on its platform amplification results with “threshold” (detectable fluorescence level) and “baseline” (defined limit of PCR cycles) parameters used to determine the Ct (cycle threshold) of each sample as shown in Figure 4. Based on our results, we propose a classification algorithm that defines Ct <34 as positive, 34<Ct>35 indicates a retest, and Ct>36 as negative. On the other hand, the GeneXpert MTB/RIF results are provided as five semiquantitative detection levels: trace, very low, low, medium, and high. Table 4 shows the correlation between the results of the Q3 Plus (positive or negative, based on the above algorithm) and the semiquantitative results provided by GeneXpert MTB/RIF. Of the 45 positive samples in GeneXpert MTB/RIF, seven negative results were presented in Q3 Plus and culture, with the semiquantitative of these samples being traced for three samples, very low for two, and low for two samples. When analyzing the agreement of molecular methods with culture, we obtained a substantial agreement (k= 0.62) for Q3 Plus and a fair agreement (k=0.31) for GeneXpert MTB/RIF, with sensitivity of 92% and 88% and specificity of 61% and 62%, respectively for each instrument. When analyzing the agreement of the molecular methods with the TB case classification, the portable protocol (FTA card and Q3-Plus) presented a moderate agreement (k=0.57) and GeneXpert MTB/RIF showed a fair agreement (k=0.32), with a sensitivity and specificity respectively of 75% and 81% and GeneXpert MTB/RIF 86% and 100%. The results obtained by both instruments, GeneXpert MTB/RIF and Q3-Plus, showed a substantial agreement (k =0.62). The positivity by all four methods and their convergence is shown in Figure 7. Of the 98 samples, 12 samples were positive by all four diagnostic techniques (culture, case TB, GeneXpert MTB/RIF, FTA card, and Q3-Plus instrument), 15 were positive by the Q3-Plus system, GeneXpert and case TB, 01 samples was positive by case TB and culture, 06 were positive by GeneXpert and case TB, 02 were positive only by case TB, and 01 sample was positive by the portable platform and case TB. Finally, all 12 samples identified as positive by culture were also positive by the portable method, and of the 34 samples identified as positive by the Q3-Plus instrument, 28 were also positive by case TB, while all 45 samples positive by the GeneXpert MTB/RIF assay were positive by case TB.

**Table 4.** Correlation between Q3 Plus and GeneXpert MTB/RIF semiquantitative results.

| GeneXpert MTB/RIF<br>Results | Q3 Plus |  |  |  |
| --- | --- | --- | --- | --- |
|  | Positive | Negative | Retest | Total |
| Negative | 7 | 30 | 16 | 53 |
| Positive trace | 1 | 3 | 2 | 6 |
| Positive very low | 1 | 1 | 3 | 5 |
| Positive low | 1 | 2 | 2 | 5 |
| Positive medium | 4 | 0 | 2 | 6 |
| Positive high | 20 | 0 | 3 | 23 |
| Total | 34 | 36 | 28 | 98 |

**Figure 7.**
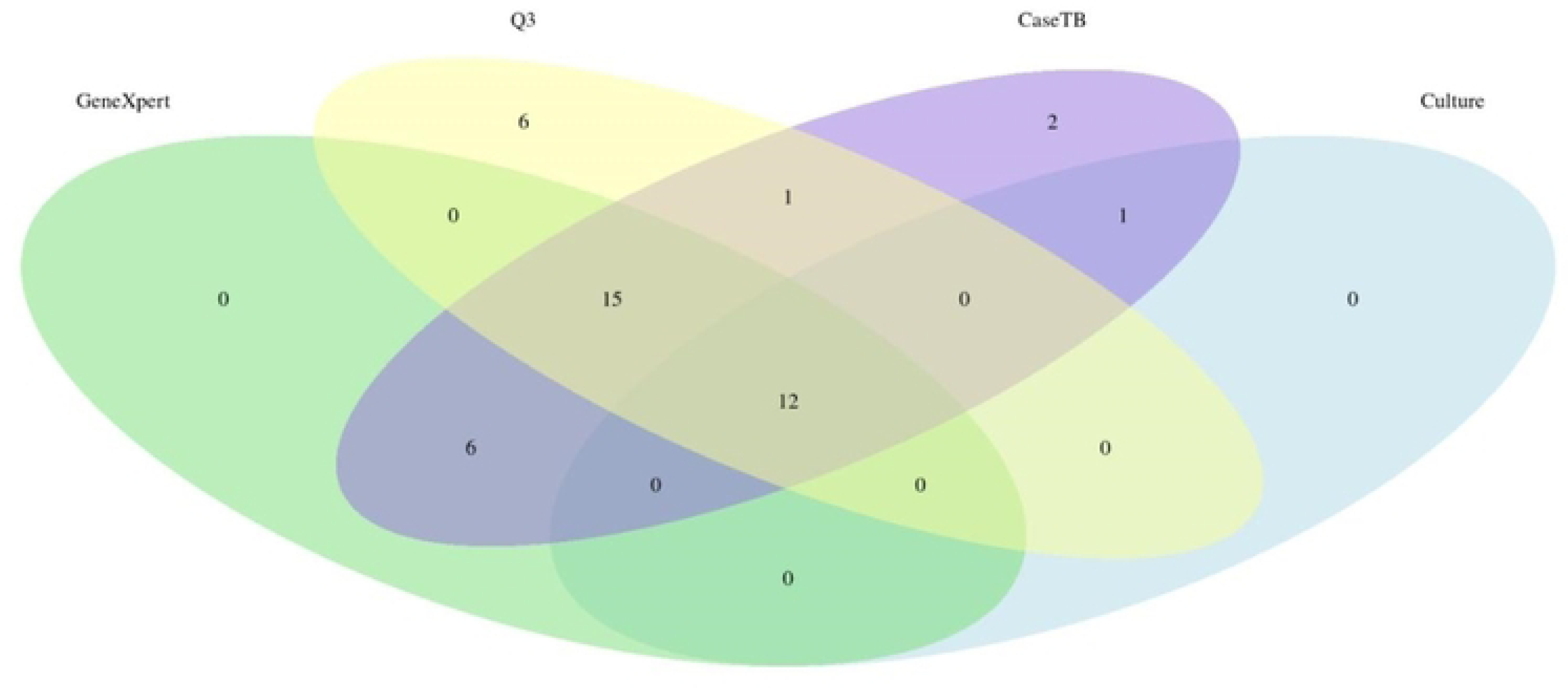
Venn diagram showing the relationship between and the number of samples that were detected by each technique. TB clinical case classification indicated 52 samples as positive, while 45 samples were detected as positive only in GeneXpert and TB Case, 28 samples were positive only in Q3 and TB case classification, 15 samples were detected simultaneously by GeneXpert MTB/RIF and culture, 17 were detected by the portable protocol and culture, and 12 samples were detected by all protocols. Out of the diagram, 46 samples were deemed negative and 28 were undetermined.

## Discussion

Tuberculosis remains a significant problem, particularly in countries with high poverty rates and areas with limited access to healthcare and infrastructure [22]. In this study, we evaluated a molecular assay to aid in the screening and detection of TB using a portable system with potential point-of-care capabilities. The use of portable platforms represents a significant advance in diagnostic technology [23]. Our study used a simplified DNA extraction protocol previously shown to liquefy the sample and to facilitate the retention of the DNA in the FTA card, which is essential for sample storage and transport [15]. FTA cards have already been used in studies with sputum, blood, and saliva samples [14,24,25,26]. Efficient nucleic acid extraction is essential for the successful diagnosis of TB by qPCR and for effectively contributing to disease control [27]. Another tool for disease control is to increase the population’s access to molecular tests, which is hampered by the difficulties of using and maintaining the technique’s sensitive instruments, such as centrifuges and thermocyclers, in areas with limited or non-existent laboratory infrastructure [28].

In the Brazilian Unified Health System (SUS), the GeneXpert MTB/RIF test is implemented for routine TB diagnosis and rifampin resistance detection [8]. Since this instrument requires a robust laboratory infrastructure, its use is impaired by the logistical difficulties of hard-to-reach areas. To overcome these obstacles, we sought to validate a simplified DNA extraction method from sputum that could be used in low infrastructure setting or point of care situations, since a portable qPCR instrument has already been developed [17].

In this study, two simple methods for DNA extraction from sputum were evaluated. The first is based on sonication, which is a common in-house method where cell membrane rupture occurs through ultrasound. The second method uses a strong denaturing chemical and proteinase K followed by solubilization by detergents embedded in the FTA Elute Micro Card to release the bacilli DNA [15,29]. The two simplified DNA extraction protocols showed an important agreement (70%), with eleven out of 29 samples testing positive by qPCR with DNA obtained by FTA card protocol, and nine out of 29 samples testing positive with DNA obtained by sonication. Three out of the 29 samples, however, showed divergence in detection by qPCR relative to the GeneXpert assay classification. Two true positives samples were detected by FTA card extraction and positive by culture and GeneXpert MTB/RIF were negative by the sonication extraction method. These divergences may occur due to the breaking of genomic DNA in the sonication process. Mandakhalikar et al. (2018) also used the sonication DNA extraction method and reported that high temperatures in the protocol can hinder DNA recovery. In addition, the cavitation in the water bath sonicator is low, making the process uneven [30]. Accordingly, a positive sample by the FTA card protocol and positive by GeneXpert MTB/RIF but negative by sonication may have been a function of sample heterogeneity or the low presence of the genomic targets (low bacterial index). Interestingly however, discordant samples #20, #22, and #28 show high Ct in the qPCR, suggesting that some of the above considerations might play a role in the discrepant results [30, 30, 31]. Using a smaller volume for elution in both protocols could potentially yield better results, but this was not evaluated. The comparative evaluation of the results of the two extraction protocols followed by qPCR in a standard instrument showed a strong agreement with the results obtained by the GeneXpert MTB/RIF assay, with disagreement between only two samples (#16 and #21, both negative by standard qPCR and positive by GeneXpert MTB/RIF). The two samples were also culture negative, suggesting that they are false positives. This is suggested by the low specificity of 79% when GeneXpert MTB/RIF was compared to the culture. Even considering a sensitivity of 100%, such lack of specificity can influence the diagnosis of negative cases, impacting clinical outcomes. It is worth noting that GeneXpert MTB/RIF, like any qPCR technique, equally amplifies DNA fragments from alive and dead bacilli (that is, viable or not for growth in culture), thus sometimes producing “false positive results” compared to culture. Indeed, Silva et al. (2019) [32] showed that patients undergoing treatment or with a history of positive TB can be qPCR positive for up to five years, highlighting the possible low specificity of such tests [32]. When the results of the sonication protocol were compared with GeneXpert MTB/RIF, there was inconsistency in four samples that were false negatives. Of these four, two were also culture positive. The relationship between the detection performance of each technique can be visualized in a Venn diagram (Figure 7).

Results obtained by the FTA card protocol and the portable qPCR instrument Q3-Plus showed perfect sensitivity (100%) and specificity (100%) when compared to culture, in agreement with previous data from our group. Ali and colleagues (2020) also used FTA Elute Micro cards, and the results showed almost perfect agreement in sensitivity and specificity when compared with GeneXpert MTB/RIF and culture, with only one false positive [15]. qPCR results obtained by the sonication protocol showed less sensitivity and specificity in relation to culture (80% and 89%). Interestingly, specificity was slightly greater than that of GeneXpert MTB/RIF. However, issues related to false positive and false negative results would require improvements in the sonication protocol to produce more accurate and acceptable results.

According to the WHO (34), greater sensitivity and low specificity are recommended in screening tests, as in the case of the portable POC platform described in the present study. Although the sonication protocol uses commonly available reagents and common laboratory equipment, it requires washing steps just like commercial kits, resulting in a longer and more laborious procedure. As for the limit of detection (LOD), it provided a good detection capacity (up to 19.3 CFU/mL). The LOD_95%_ obtained in our study was similar to that of Chakravorty et al. (2017), who analyzed the performance of the GeneXpert MTB/RIF Ultra as a point-of-care (POC) assay suitable for diagnosing TB and obtained a LOD_95%_ of 15.6 CFU/mL. The FTA card protocol, however, showed greater sensitivity and specificity, as well as other benefits such as easiness of operation (a lesser requirement for laboratory equipment, for example), transportation, storage, and conservation of the genetic material [33]. Evaluation of the DNA amplification with the Q3-Plus system showed fair agreement (Kappa = 0.62) with the GeneXpert/MTB RIF assay, which was more sensitive than GeneXpert when compared to culture. Despite both methods being molecular, the Q3-Plus detected more cases than GeneXpert MTB/RIF. Despite the high sensitivity in our study, the specificity was low, which could be justified by the medical history of the patients, where some were already in treatment, consistent with the studies by Zhang et al. 2021 and Silva et al. 2017 [32,35]. Zhang and colleagues, as well as Silva and colleagues, highlighted in their studies that the decrease in specificity is related to the previous history of TB, and molecular methods can detect patients in treatment, i.e., with dead bacilli, justifying the low specificity (61% in our study with 22 false positives). It is known that only live bacilli grow in culture, and these studies also emphasize the importance of knowing the patient’s history, especially if they are in treatment, along with the molecular diagnosis [32,35]. Our results provide evidence, however, that the simplified DNA extraction protocol followed by qPCR detection in the Q3-Plus system can detect *M. tuberculosis* DNA at all stages of the disease.

In our study, the GeneXpert MTB/RIF assay showed fair agreement with 30 false positives and a specificity of 62% when compared to culture, the gold standard. Although the GeneXpert MTB/RIF assay showed better results and a faster turnaround time of 80– 90 minutes, it is expensive (in Brazil, approximately US$10 after up to 90% subsidies) and is not available in areas with limited infrastructure and remote regions. In contrast, the portable Q3-Plus system uses a smaller reaction volume (only 5 μl) and provides results in up to 4 hours. Additionally, the portable platform can be transported to remote health units or regions where the only available infrastructure is electricity and a roof, thus aiding in the diagnosis of TB and effectively helping to contain the spread of the disease in local communities.

## Conclusion

The present study validated a simplified and portable procedure for qPCR screening of individuals suspected of TB infection, suitable for use in point-of-care, low-infrastructure settings or in areas where people are highly vulnerable and lack access to healthcare centers, thus effectively making molecular diagnosis accessible to everyone.

## Data Availability

All data produced in the present work are contained in the manuscript.

## Acknowledgements

We are greatful to the Department of Tisiology and Leprosy of the Health Department at the University Hospital of Canoas (Canoas, Brazil), for their support in obtaining the samples.

## Funding

This work was funded by grant CNPq 445954/2020-5 from Conselho Nacional de Desenvolvimento Científico e Tecnológico (CNPq, https://www.gov.br/cnpq/pt-br) to ADTC and INCT-TB FAPERGS 17/1265 from CNPq and Fundação de Amparo à Pesquisa do Estado do Rio Grande do Sul (FAPERGS, https://fapergs.rs.gov.br/inicial) to MLRR. TSS is a fellowship holder at CAPES (Fundação Coordenação de Aperfeiçoamento de Pessoal de Nível Superior, https://www.gov.br/capes/pt-br). ADTC is a CNPq Productivity Fellow (level 2). The funders played no role in the study design, data collection and analysis, decision to publish, or preparation of the manuscript.

## Statements

### Ethics

This research was approved by the Ethics Committee of ULBRA (CAAE 0697116.7.0000.5349 to GLB) and by the Ethics Committee of FIOCRUZ (CAAE 4423120.0.1001.5248 to ADTC).

### Consent for publication

Written informed consent for publication was obtained from all individual participants included in the study.

### Conflict of interest

The authors declare no conflict of interest exists.

## Notes

### Competing Interest Statement

The authors have declared no competing interest.

### Funding Statement

This work was funded by grant CNPq 445954/2020-5 from Conselho Nacional de Desenvolvimento Cientifico e Tecnologico (CNPq, https://www.gov.br/cnpq/pt-br) to ADTC and INCT-TB FAPERGS 17/1265 from CNPq and Fundacao de Amparo a Pesquisa do Estado do Rio Grande do Sul (FAPERGS, https://fapergs.rs.gov.br/inicial) to MLRR. TSS is a fellowship holder at CAPES (Fundacao Coordenacao de Aperfeicoamento de Pessoal de Nivel Superior, https://www.gov.br/capes/pt-br). ADTC is a CNPq Productivity Fellow (level 2).

### Author Declarations

This research was approved by the Ethics Committee of Universidade Luterana do Brasil (0697116.7.0000.5349 to GLB) and by the Ethics Committee of Fundacao Oswaldo Cruz (CAAE 4423120.0.1001.5248 to ADTC)

## References

1. Boletim Epidemiológico de Tuberculose - Número Especial [Internet]. Gov.br. [cited 2023 Jul 3]. Available from: https://www.gov.br/saude/pt-br/centrais-de-conteudo/publicacoes/boletins/epidemiologicos/especiais/2023/boletim-epidemiologico-de-tuberculose-numero-especial-mar.2023/view

2. World Health Organization. (2022). Global tuberculosis report 2022. World Health Organization. https://apps.who.int/iris/handle/10665/363752. License: CC BY-NC-SA 3.0 IGO.

3. Rasool G, Khan AM, Mohy-Ud-Din R, Riaz M. Detection of mycobacterium tuberculosis in afb smear-negative sputum specimens through MTB culture and GeneXpert® MTB/RIF assay. Int J Immunopathol Pharmacol. 2019;33. doi:10.1177/2058738419827174

4. Brasil (2011) Ministério da Saúde. Secretária de Vigilância em Saúde. Departamento de vigilância epidemiológica. Manual de ecomendações para controle da tuberculose no Brasil. ed 1, 284 p 2011 ISBN 978-85-334-1816-5

5. Bento J, Silva AS, Rodrigues F, Duarte R. Métodos diagnósticos em tuberculose. Acta Med Port., 2011; 24(1): 145–154

6. Lima TM de Belotti NCU, Nardi SMT, Pedro H da SP. Teste rápido molecular GeneXpert MTB/RIF para diagnóstico da tuberculose. Rev Panamazonica Saude. 2017;8(2):65–76. doi:10.5123/S2176-62232017000200008

7. Ullah I, Shah AA, Basit A, et al. Rifampicin resistance mutations in the 81 bp RRDR of rpoB gene in Mycobacterium tuberculosis clinical isolates using Xpert MTB/RIF in Khyber Pakhtunkhwa, Pakistan: A retrospective study. BMC Infect Dis. 2016;16(1). doi:10.1186/s12879-016-1745-2

8. Opota O, Zakham F, Mazza-Stalder J, Nicod L, Greub G, Jaton K. Added value of Xpert MTB/RIF Ultra for diagnosis of pulmonary tuberculosis in a low-prevalence setting. J Clin Microbiol. 2019;57(2). doi:10.1128/JCM.01717-18

9. Ali N, Rampazzo RDCP, Costa ADiT, Krieger MA. Current Nucleic Acid Extraction Methods and Their Implications to Point-of-Care Diagnostics. Biomed Res Int. 2017;2017. doi:10.1155/2017/9306564

10. Ndhlovu V, Mandala W, Sloan D, Kamdolozi M, Caws M, Davies G. Evaluation of the efficacy of two methods for direct extraction of DNA from Mycobacterium tuberculosis sputum. J Infect Dev Ctries. 2018;12(12):1067–1072. doi:10.3855/jidc.10592

11. Amaro A, Duarte E, Amado A, Ferronha H, Botelho A. Comparison of three DNA extraction methods for Mycobacterium bovis, Mycobacterium tuberculosis and Mycobacterium avium subsp. avium. Lett Appl Microbiol. 2008;47(1):8–11. doi:10.1111/j.1472-765X.2008.02372

12. Landis JR, Koch GG. The measurement of observer agreement for categorical data. Biometrics. 1977;33(1):159–174.

13. Kolia-Diafouka P, Godreuil S, Bourdin A, et al. Optimized Lysis-Extraction Method Combined with IS6110-Amplification for Detection of Mycobacterium tuberculosis in Paucibacillary Sputum Specimens. Front Microbiol. 2018;9. doi:10.3389/fmicb.2018.02224

14. Kang M, Yang JS, Kim Y, Kim K, Choi H, Lee SH. Comparison of DNA extraction methods for drug susceptibility testing by allele-specific primer extension on a microsphere-based platform: Chelex-100 (in-house and commercialized) and MagPurix TB DNA Extraction Kit. J Microbiol Methods. 2018;152. doi: 10.1016/j.mimet.2018.07.019

15. Ali N, Bello GL, Rossetti MLR, Krieger MA, Costa ADT. Demonstration of a fast and easy sample-to-answer protocol for tuberculosis screening in point-of-care settings: A proof of concept study. PLoS One. 2020;15(12 December). doi: 10.1371/journal.pone.0242408

16. Rabodoarivelo MS, Brandao A, Novella MCC, et al. Detection of multidrug-resistant tuberculosis from stored DNA Samples: A multicenter study. Int J Mycobacteriol. 2018;7(1). doi: 10.4103/ijmy.ijmy_193_17

17. Cereda M, Cocci A, Cucchi D, Raia L, Pirola D, Bruno L, et al. Q3: A compact device for quick, high precision qPCR. Sensors (Basel).;18(8):2583.

18. Case definitions. Genebra, Switzerland: World Health Organization; 2010

19. Brasil Ministério da Saúde. Manual nacional de vigilância laboratorial da tuberculose e outras micobactérias. Brasília: Ministério da Saúde; 2008.

20. Bello GL, Morais FCL, Wolf JM, et al. Improvement of Mycobacterium tuberculosis detection in sputum using DNA extracted by sonication. Brazilian Journal of Infectious Diseases. 2020;24(5). doi: 10.1016/j.bjid.2020.08.006

21. 21. Schimid KB (2014) Comparação de duas metodologias moleculares para diagnóstico de tuberculose. Dissertation, Universidade Federal do Rio Grande do Sul.

22. Palomino JC, Martin A, Von Groll A, Portaels F. Rapid culture-based methods for drug-resistance detection in Mycobacterium tuberculosis. J Microbiol Methods. 2008 Oct;75(2):161–6. doi: 10.1016/j.mimet.2008.06.015.

23. Rampazzo RCP, Graziani AC, Leite KK, Surdi JA, Biondo CA, Costa MLN, Jacomasso T, Cereda M, De Fazio M, Bianchessi MA, Moreira OC, Britto C, Costa JDN, Góes VM, da Silva AJ, Krieger MA, Costa ADT. Proof of Concept for a Portable Platform for Molecular Diagnosis of Tropical Diseases: On-Chip Ready-to-Use Real-Time Quantitative PCR for Detection of Trypanosoma cruzi or Plasmodium spp. J Mol Diagn. 2019 Sep;21(5):839–851. doi: 10.1016/j.jmoldx.2019.04.008

24. Dippenaar A, Ismail N, Grobbelaar M, Oostvogels S, de Vos M, Streicher EM, Heupink TH, van Rie A, Warren RM. Optimizing liquefaction and decontamination of sputum for DNA extraction from Mycobacterium tuberculosis. Tuberculosis (Edinb). 2022 Jan; 132:102159. doi: 10.1016/j.tube.2021.102159.

25. de Vargas Wolfgramm E, de Carvalho FM, da Costa Aguiar VR, De Nadai Sartori MP, Hirschfeld-Campolongo GCR, Tsutsumida WM, et al. Simplified buccal DNA extraction with FTA®Elute Cards. Forensic Sci Int Genet. 2009;3: 125–127.

26. Petry IM dos S, Rolim F dos S, Soares T dos S, Carvalho MER de Prophiro JS, Leote DS, Rossetti ML. Detecção de Leishmania spp. em soro canino fixado em cartão. Research, Society and Development, v. 11, n. 16, e582111638434, 2022.

27. Leung ETY, Zheng L, Wong RYK, et al. Rapid and simultaneous detection of Mycobacterium tuberculosis complex and Beijing/W Genotype in sputum by an optimized DNA extraction protocol and a novel multiplex real-time PCR. J Clin Microbiol. 2011;49(7). doi:10.1128/JCM.00108-11

28. Ahmad R, Xie L, Pyle M, et al. Erratum: A rapid triage test for active pulmonary tuberculosis in adult patients with persistent cough. Sci Transl Med. 2019;11(516). doi:10.1126/scitranslmed. aaz9925

29. Ali N, Rampazzo RDCP, Costa ADiT, Krieger MA. Current Nucleic Acid Extraction Methods and Their Implications to Point-of-Care Diagnostics. Biomed Res Int. 2017;2017. doi:10.1155/2017/9306564

30. Mandakhalikar KD, Rahmat JN, Chiong E, Neoh KG, Shen L, Tambyah PA. Extraction and quantification of biofilm bacteria: Method optimized for urinary catheters. Sci Rep. 2018;8(1):8069. doi:10.1038/s41598-018-26342-3

31. Cox AP, Tosas O, Tilley A, et al. Constraints to estimating the prevalence of trypanosome infections in East African zebu cattle. Parasit Vectors. 2010;3(1). doi:10.1186/1756-3305-3-82

32. Silva TMD, Soares VM, Ramos MG, Santos AD. Accuracy of a rapid molecular test for tuberculosis in sputum samples, bronchoalveolar lavage fluid, and tracheal aspirate obtained from patients with suspected pulmonary tuberculosis at a tertiary referral hospital. J Bras Pneumol. 2019;45(2): e20170451.doi:10.1590/1806-3713/e20170451

33. Da Cunha Santos G. FTA cards for preservation of nucleic acids for molecular assays a review on the use of cytologic/tissue samples. In: Archives of Pathology and Laboratory Medicine. Vol 142.; 2018. doi:10.5858/arpa.2017-0303-RA

34. Jul 17. WHO consolidated guidelines on tuberculosis Module 2: Screening – Systematic screening for tuberculosis disease. Pan American Health Organization (PAHO). Available from: https://www.paho.org/en/documents/who-consolidated-guidelines-tuberculosis-module-2-screening-systematic-screening

35. Zhang, Z., Du, J., Liu, T., Wang, F., Jia, J., Dong, L., Zhao, L., Xue, Y., Jiang, G., Yu, X., & Huang, H. (2021). EasyNAT MTC assay: A simple, rapid, and low-cost cross-priming amplification method for the detection of mycobacterium tuberculosis suitable for point-of-care testing. Emerging Microbes and Infections, 10(1). 10.1080/22221751.2021.1959271

